# The Age Pattern of the Male- to- Female Ratio in Mortality from COVID-19 Mirrors that of Cardiovascular Disease but not Cancer in the General Population

**DOI:** 10.1101/2020.07.10.20149013

**Authors:** Ila Nimgaonkar, Linda Valeri, Ezra Susser, Sabiha Hussain, Jag Sunderram, Abraham Aviv

## Abstract

**Background:** Males are at a higher risk of dying from COVID-19. Older age and cardiovascular disease are also associated with COVID-19 mortality. We compared the male-to-female (sex) ratios in mortality by age for COVID-19 with cardiovascular mortality and cancer mortality in the general population.

**Methods:** We obtained data from official government sources in the US and five European countries: Italy, Spain, France, Germany, and the Netherlands. We analyzed COVID-19 deaths by sex and age in these countries and similarly analyzed their deaths from cardiovascular disease (coronary heart disease or stroke) and cancer, the two leading age-related causes of death in middle-to-high income countries.

**Findings:** In both the US and European countries, the sex ratio of deaths from COVID-19 exceeded one throughout adult life. The sex ratio increased up to a peak in midlife, and then declined markedly in later life. This pattern was also observed for the sex ratio of deaths from cardiovascular disease, but not cancer, in the general populations of the US and European countries.

**Interpretation:** The sex ratios of deaths from COVID-19 and from cardiovascular disease exhibit similar patterns across the adult life course. The underlying mechanisms are poorly understood, but could stem partially from sex-related biological differences that underlie the similar pattern for cardiovascular disease. These include, we propose, comparatively longer telomeres in females, ovarian hormones, and X chromosome mosaicism.

**Funding:** The authors received no specific funding for this work.

**Research in Context:** *Evidence before this study:* Mortality from COVID-19 is higher in males and older persons. We searched PubMed.gov in June 2020, with no date restrictions, for articles published in English using the search terms “COVID-19”, “deaths”, “mortality”, “sex”, “male”, “female”, “age”, “disaggregated”, “ratio”, and “stratified”. We identified studies in several countries that stratified COVID-19 mortality data by age or by sex, but no study examined male-to-female (sex) ratios by age at a multi-national level.

*Added value of this study:* To our knowledge, this is the first study to aggregate data on COVID-19 deaths at a multi-national level, and analyze how the sex ratio in deaths varies by age, taking into account the population at risk in each sex/age stratum. We found a distinctive pattern in sex ratio of deaths by age, illuminating the nature of the sex effect on death from COVID-19. Moreover, we found a similar pattern in sex ratio of deaths by age for cardiovascular disease, which is strongly associated with increased risk of dying from COVID-19. We did not find a similar age pattern for the sex ratio in deaths by cancer.

*Implications of all the available evidence:* The sex ratio in deaths from COVID-19 peaks in middle age and decreases at older ages, a pattern mirrored in deaths from cardiovascular disease. This intriguing similarity warrants research about whether sex-based differences in mortality from COVID-19 and from cardiovascular disease in general are partly due to common biological causes.

## Introduction

More males than females die from COVID-19, the disease caused by the SARS-CoV-2 virus^1^. This was first observed in China, where 64% of COVID-19 deaths occurred in males^1^. As the epidemic spread worldwide, other countries similarly observed a higher percentage of deaths from COVID-19 occurring in males^1^. The higher number of deaths among males is consistent with mortality patterns observed in several major viral epidemics/pandemics of the 20^th^ and 21^st^ centuries, including the Western African Ebola virus epidemic (2013-2016)^2^ and the H1N1 Spanish Flu pandemic of 1918^3^. Adult men also have an overall higher mortality rate than adult women from seasonal influenza based on an analysis of data in the US between 1997-2007, with some variation depending on age group and underlying conditions^4^.

In contrast, COVID mortality by age differs from other viral pandemics. More than 80% of COVID-19 deaths in the US and European countries have occurred in individuals older than 65 years^5^, with very few deaths in young children^6^. Seasonal influenza causes relatively more pediatric deaths, especially in infants under the age of six months^7^, in addition to a disproportionate number of deaths in individuals over the age of 65 years^8^. Several major viral pandemics of the 20^th^ and 21^st^ centuries have also shown different age-based mortality patterns from COVID-19. For instance, in the Spanish Flu of 1918 a large proportion of deaths were in young adults^9^, and in the 2009 H1N1 influenza pandemic a large proportion occurred in children and non-elderly adults^10^. Therefore, COVID-19 mortality trends are consistent with sex-based, but not with age-based patterns seen in many other viral pandemics.

In searching for explanations for the distinctive pattern of deaths in COVID-19, we first examined variation by age group in the male-to-female (sex) ratio of mortality in data from the US and five European countries. Given that cardiovascular disease (CVD) has been strongly associated with increased risk of mortality in COVID-19^11^, we next examined variation in the sex ratio of CVD mortality by age group in the same countries. As CVD and cancer are the two major age-related disease categories that largely determine survival of adults in middle-and-high income societies, we further examined variation by age in the sex ratio of cancer mortality. Here we report our findings and offer potential explanations regarding their meaning.

## Methods

We focused our analysis on the US, Italy, Spain, France, Germany, and the Netherlands because of (a) availability of their sex- and age-disaggregated mortality data from COVID-19, (b) data stratification into age group bins of 10 years or less, and (c) a high number of cumulative deaths from COVID-19 (sources are shown in Supplementary Table 1). COVID-19 mortality data were retrieved from national databases including the US Centers for Disease Control, the Italian National Institute of Health, the French Institute for Demographic Studies, the Spanish Ministry of Health, the German Federal Ministry of Health, and the Dutch Ministry for Health, Welfare and Sport. The data were retrieved on June 18^th^, 2020 and reflect the cumulative COVID-19 deaths in each country from the beginning of the pandemic up to dates between May 29^th^ – June 17^th^ 2020 (see Supplementary Table 1 and Supplementary Table 2 for details). In the US, data were included for age groups 25 years and older, and stratified into 10-year bins. For the European Countries, data were included for age groups 30 years and above and stratified into 10-year bins, and deaths were combined across the five countries by age group and sex. Age groups with less than 200 total deaths were excluded from the analysis. The population at risk in each age group was retrieved from populationpyramid.net, a website that aggregates data from the United Nations Department of Economic and Social Affairs, Population Division.

For the analysis of mortality from CVD and cancer, data were extracted from the World Health Organization Mortality Database (https://www.who.int/healthinfo/mortality_data/en/), which collects national data on deaths from civil registries. The Database contains number of deaths by country, year, sex, age group and cause of death, and population size by country, year, sex and age group. The causes of death are categorized by International Classification of Disease (ICD)-10 codes. CVD deaths principally included deaths from coronary heart disease (ICD-10 codes I20-I25) or stroke (I60-I69)^12^. Cancer death data included deaths from all neoplasms (C00-C97, D00-D48) (Supplementary Table 3). Data was extracted for France (2014), Germany (2015), Italy (2015), Netherlands (2016), Spain (2015), and the US (2015), with the year of the most recent data available indicated in parentheses (Supplementary Table 4 and Supplementary Table 5). Population adjustment was done using population data from the same years as the mortality data. These population data were also extracted from the World Health Organization Mortality Database.

Plots and data visualizations were created using the ggplot2 package in R (https://ggplot2.tidyverse.org).

## Results

Stratified by age, and adjusted for population at risk in each age group (Supplementary Figure 1, Supplementary Table 2), the sex ratios for COVID-19 deaths among adults in the US and five European countries (Italy, Spain, France, Germany, and the Netherlands) showed similar overall patterns, initially rising to a peak in midlife and then falling (Figure 1). Specifically, the sex ratio of mortality peaked between the ages of 35-44 years in the US and 60-69 years in the European countries, and then progressively declined at older ages without dropping below one.

**Figure 1.**
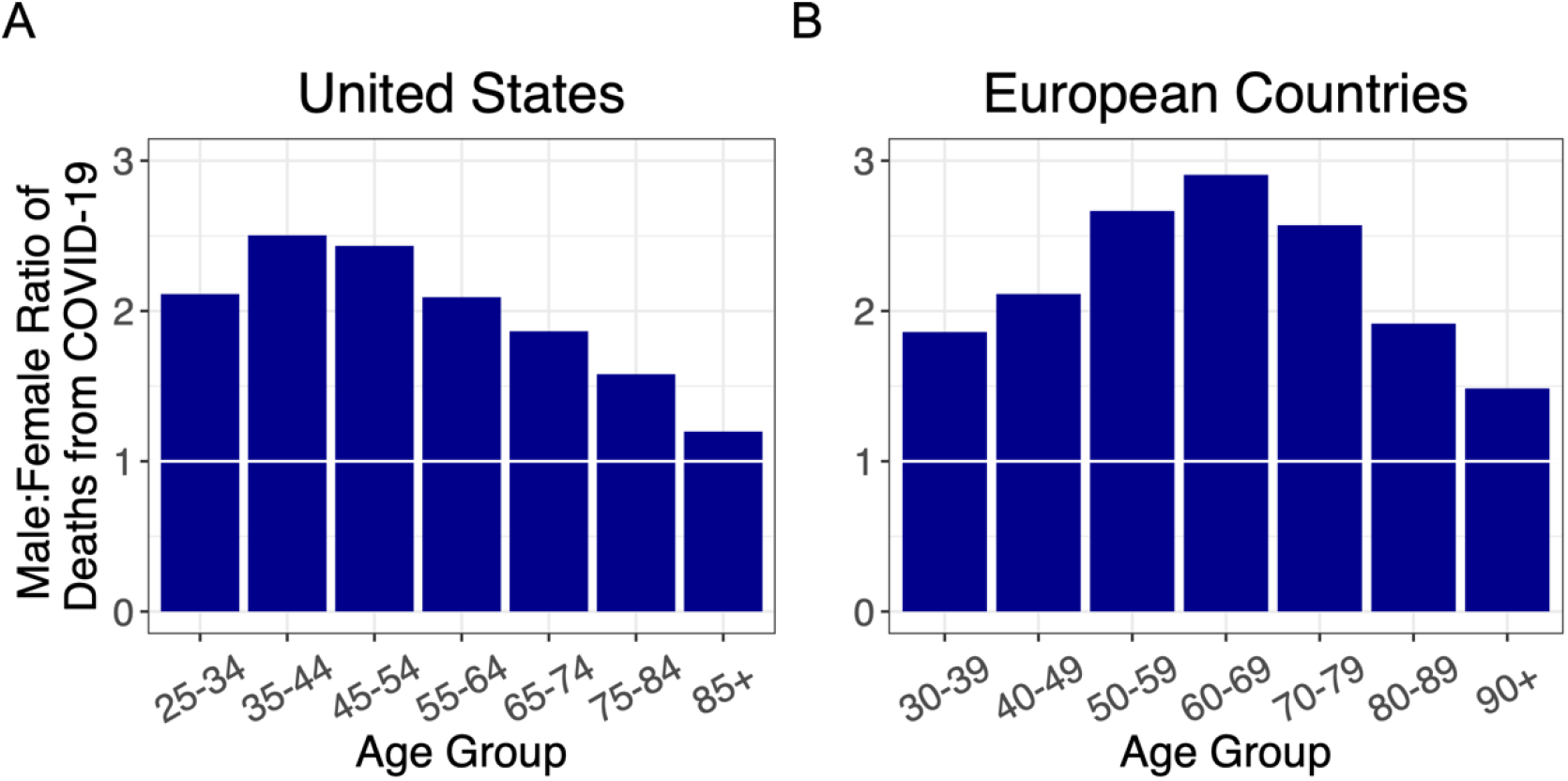
Ratios of male to female deaths from COVID-19 (adjusted for population sex distribution) for (A) the US, and (B) combined ratios for five European countries: Italy, France, Spain, Germany, and the Netherlands. A 1:1 ratio is indicated by white markers.

We next examined the sex ratio of CVD mortality by age in these countries (Figure 2). The sex ratio for CVD mortality initially rose to a peak in midlife and then declined, in both the US and Europe. For ages younger than 70 years, the sex ratio for CVD mortality in Europe was higher than in the US.

**Figure 2.**
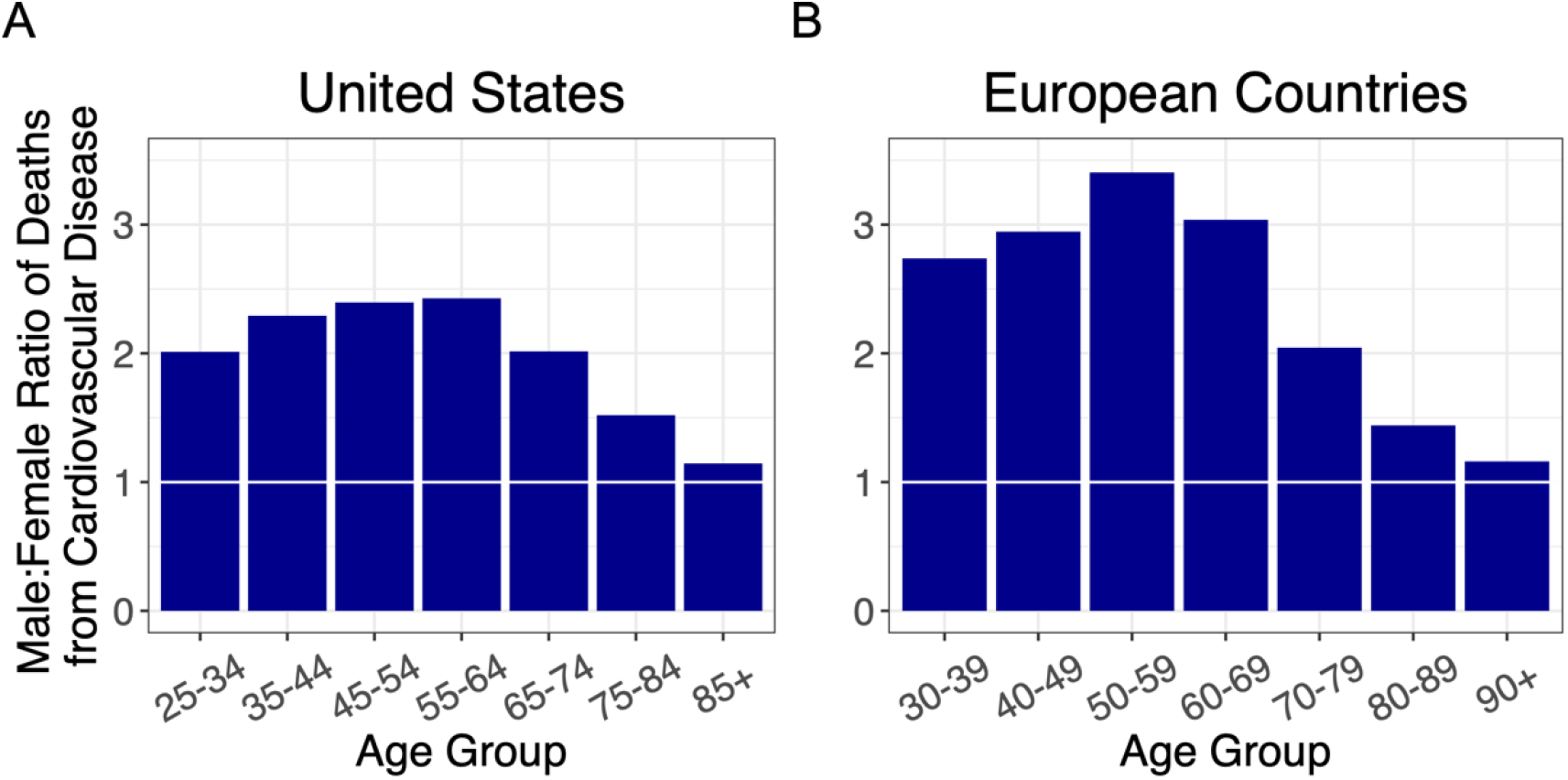
Ratios of male to female deaths from cardiovascular disease (adjusted for population sex distribution) for (A) the US, and (B) combined ratios for the European countries: Italy, France, Spain, Germany, and the Netherlands. A 1:1 ratio is indicated by white markers.

As cancer is the other leading cause of adult mortality in the US and Europe^13^, we also examined the sex ratio for cancer mortality by age (Figure 3). The sex ratio for cancer mortality increased after midlife with no evidence of decline thereafter.

**Figure 3.**
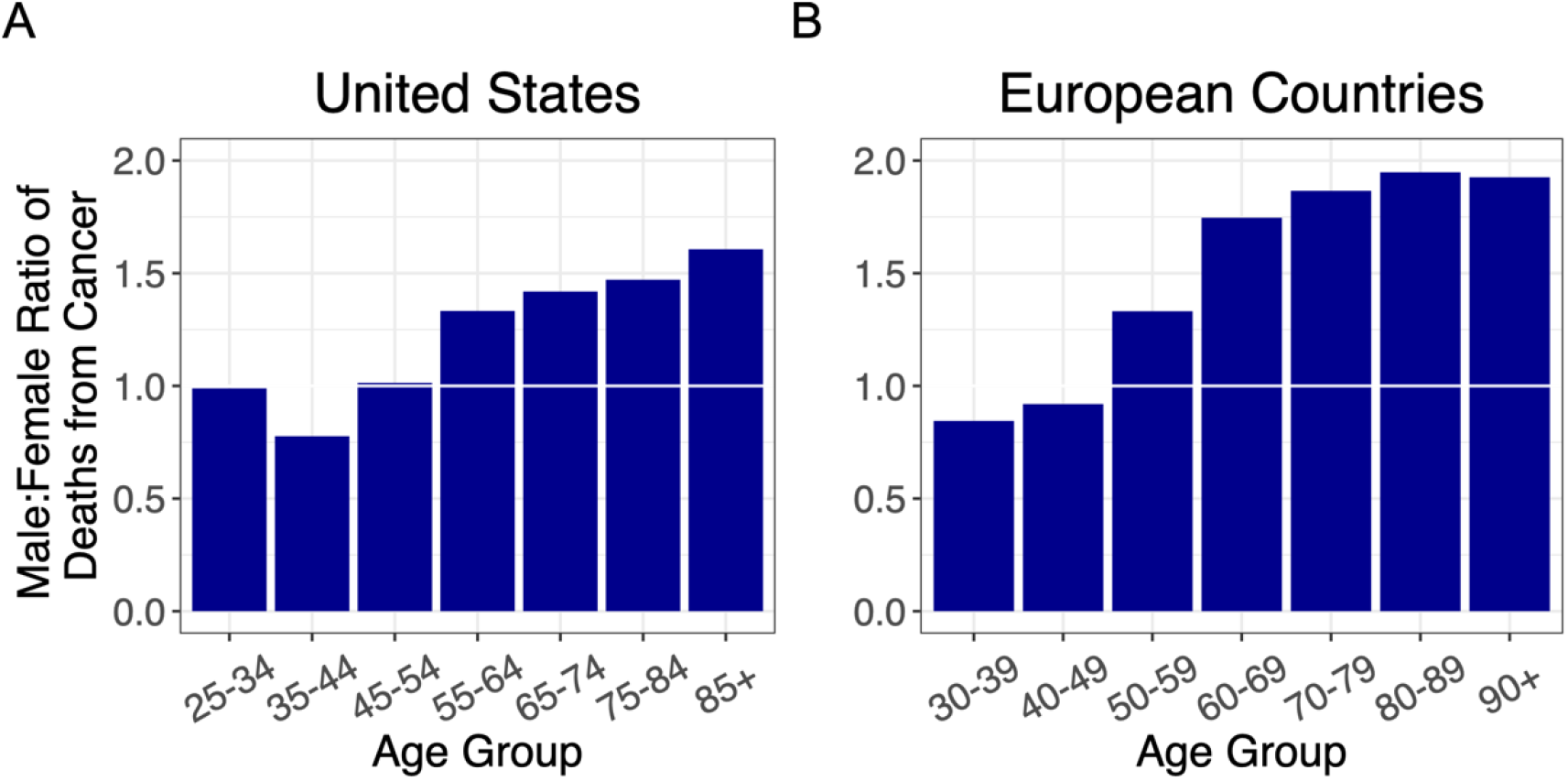
Ratios of male to female deaths from cancer (adjusted for population sex distribution) for (A) the US, and (B) combined ratios for the European countries: Italy, France, Spain, Germany, and the Netherlands. A 1:1 ratio is indicated by white markers.

When the sex ratios of mortality by age for COVID-19, CVD and cancer were overlaid, the COVID-19 profile mirrored that of CVD, although, the ratio for CVD was much higher in Europe than the US for ages younger than 70 years (Figure 4). The profile of the sex mortality ratio by age for cancer, however, clearly differed from the ratios for COVID-19 and for CVD.

**Figure 4.**
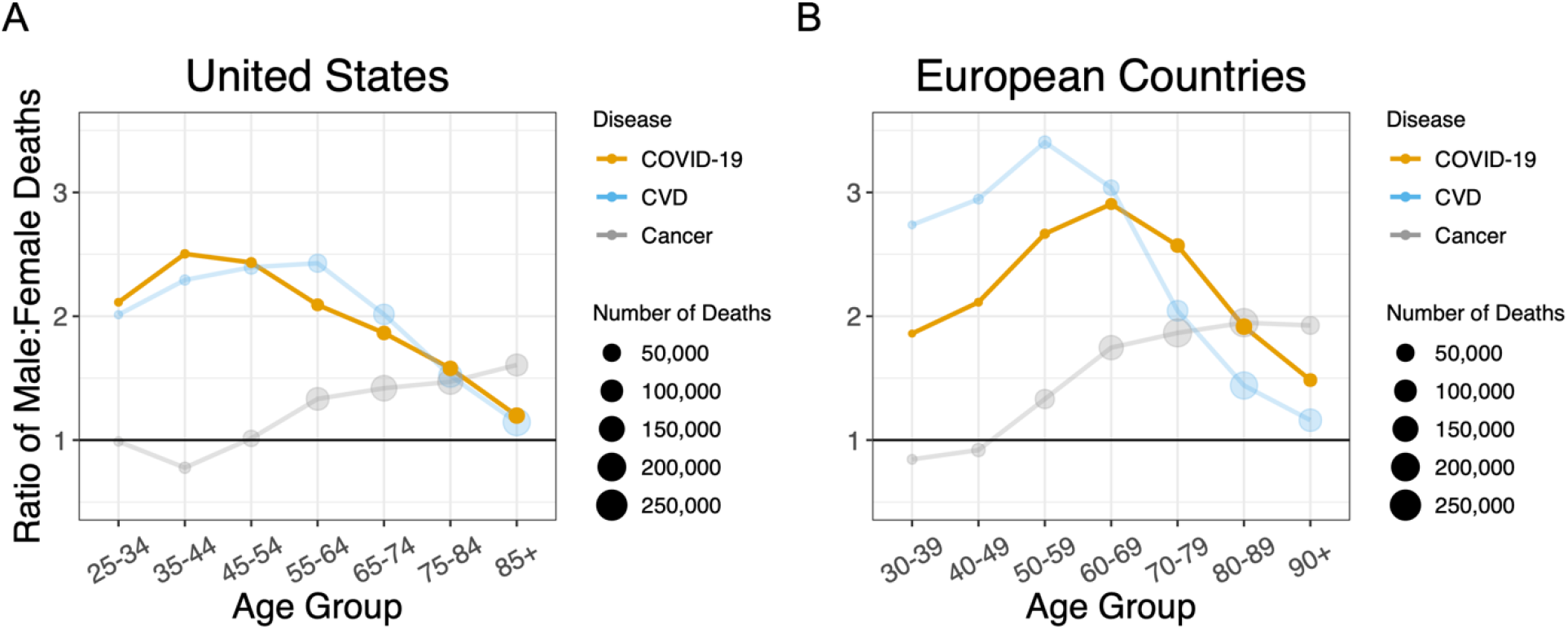
Sex ratios by age group in deaths for COVID-19, cardiovascular disease, and cancer in (A) the US and (B) the European Countries. A 1:1 ratio is indicated by black line markers. Ratios are adjusted for sex distribution in the population at different age groups.

## Discussion

### General Considerations

Our key findings are as follows: (i) for all age groups, the death rate from COVID-19 was higher in males than females; (ii) the sex ratio rose to a peak in midlife and then narrowed with increasing age; and (iii) the sex mortality ratio for COVID-19 by age mirrored that for CVD but not cancer mortality. Since the COVID-19 data were not linked to population databases with individual-level health information, we could not specifically examine the potential contribution of CVD to the age pattern of the sex mortality ratio from COVID-19. We propose, nonetheless, based on data displayed in Figure 4 that the sex ratio for COVID-19 mortality, particularly for persons older than 60 years, might partially reflect the same underlying mechanisms that drive the age pattern of the sex mortality ratio for CVD in the general population.

What then might be the mechanisms explaining the similar age patterning of the sex ratio for mortality in COVID-19 and in CVD? The underlying biological reasons are likely multifactorial and complex. We review several potential explanations: telomeres, ovarian hormones, and X chromosome mosaicism.

### Telomeres

Single nucleotide polymorphisms associated with leukocyte telomere length have been used as instrumental variables in Mendelian randomization studies to infer a causal role of short telomeres in CVD and long telomeres in major cancers^14^. Older persons with short leukocyte telomere length show a higher risk of dying from CVD^15^, and a recent paper proposed that short telomere length might be a common denominator that partially explains increased mortality from COVID-19 of older persons, males and individuals with CVD^16^. Telomeres are longer in females than males from birth onwards^17,18^, a finding that might contribute to the skewed sex ratio in mortality from COVID-19 and CVD not only in older persons but also throughout the adult life course.

### Ovarian hormones

Endogenous estrogens have been associated with a protective effect against CVD in premenopausal women^19^, while predisposing women to certain cancers^20^. Higher estrogen^21^ and progesterone^22^ levels in premenopausal women might also modulate immune responses and attenuate the severity of COVID-19. In this regard, data from the US show a decline in the sex ratio of COVID-19 mortality after ages 45-54 years, which coincides with the average age that women undergo menopause^23^. However, the data from the European countries suggest otherwise, with the drop in the sex ratio of COVID-19 mortality starting in the 70-79-year age group; well beyond the average age of menopause. Clinical trials are currently investigating whether estrogen or progesterone treatment can alleviate COVID-19 symptoms, which may provide clarity to the role of ovarian hormones in COVID-19 pathogenesis (ClinicalTrials.gov identifiers NCT04359329, NCT04365127).

### X chromosome mosaicism

The two X chromosomes provide an advantage related to X-linked recessive diseases and other deleterious mutations on the X chromosome. Random inactivation *in utero* of one X chromosome in each somatic cell engenders mosaicism that provides females with somatic cell diversity and the potential for selection of cells with an X chromosome harboring advantageous variant genes^24^. X chromosome mosaicism might be particularly advantageous for surviving infectious disease, since the X chromosome encodes a number of genes engaged in immune function – therefore having two copies of these genes confers additional immunological diversity in women^25^. In addition, the X chromosome harbors *ACE2*, the gene encoding angiotensin-converting enzyme 2, the cellular receptor for SARS-CoV-2^26^. *ACE2* variants might play a role in left ventricular hypertrophy that is often the outcome of hypertension^27^, a comorbidity that tracks with age^28^. Some of these variants might also influence CVD in general^29^, the severity of COVID-19^26^, or interact with other genes and environmental factors that influence mortality from both diseases.

### Limitations

Limitations to this study include: (a) potential differences in reporting COVID-19 deaths between the countries analyzed; (b) the lack of linked individual-level health and social databases; and (c) no information on the sex ratio of survival rates among people who acquired COVID-19. The latter would require, at a minimum, COVID-19 infection rates by age and sex in the general population (including asymptomatic infections) in order to determine whether there are sex- and age-based differences in survival from it.

Additionally, the study does not account for some anomalies in the overall similar patterns for COVID-19 and CVD in the United States versus Europe. The peak sex ratio for COVID-19 occurs at an earlier age in the United States than Europe, and the sex ratio for CVD mortality is higher for Europe than the United States until the older age groups. Finally, we could not rule out that finding the lowest sex ratios in COVID-19 and CVD for the oldest age groups was partly due to selection by survival in that males who survive to an exceptionally old age are “escapers” who hardly represent the general population of older males, whereas females who survive to such an old age might be “delayers” who largely represent the general population of older females^30^.

### Conclusions

Our analyses show similar trends in the sex ratio by age of mortality from COVID-19 and from CVD. We propose that these findings might be due to some shared underlying biological mechanisms. Individual-level data are essential to examine potential shared mechanisms and to establish the potential contribution of CVD to the age patterning of the sex ratio of mortality in COVID-19. We suggest that this line of research should be pursued and might uncover some of the causal mechanisms underlying COVID-19 mortality.

## Data Availability

The data that support the findings of this study are all available in the public domain.
The COVID-19 mortality data were derived from the following sources:
US Centers for Disease Control (https://data.cdc.gov/)
Italian National Institute of Health (https://www.epicentro.iss.it/)
French Institute for Demographic Studies (https://dc-covid.site.ined.fr/)
Spanish Ministry of Health (https://www.mscbs.gob.es/)
German Federal Ministry of Health (https://www.rki.de/)
Dutch Ministry for Health, Welfare and Sport (https://www.rivm.nl/)
Additional public databases used in the study were the WHO Mortality Database (https://www.who.int/healthinfo/mortality_data/en/) and https://www.populationpyramid.net.

https://www.epicentro.iss.it/

https://dc-covid.site.ined.fr/

https://www.mscbs.gob.es/

https://www.rki.de/

https://www.rivm.nl/

https://www.who.int/healthinfo/mortality_data/en/

https://www.populationpyramid.net

## Author Contributions

I.N., L.V., E.S. and A.A. provided substantial contributions to the design of the study. I.N. and A.A. wrote the first draft of the manuscript. All coauthors provided substantial contributions to the analysis and interpretation of data, critically reviewed the manuscript for important intellectual content, provided final approval of the version to be published, and agree to be accountable for all aspects of the work presented.

## Declaration of Interests

We declare no competing interests.

## Notes

### Competing Interest Statement

The authors have declared no competing interest.

### Author Declarations

Not applicable. The data in this study were retrieved from public databases.

## References

1. GlobalHealth5050. COVID-19 sex-disaggregated data tracker. London. https://globalhealth5050.org/covid19/sex-disaggregated-data-tracker/: Global Health 50/50; 2020.

2. Agua-Agum J, Ariyarajah A, Blake IM, et al. Ebola Virus Disease among Male and Female Persons in West Africa. N Engl J Med 2016;374(1):96-8. (In eng). DOI: 10.1056/NEJMc1510305.

3. Noymer A, Garenne M. The 1918 influenza epidemic’s effects on sex differentials in mortality in the United States. Popul Dev Rev 2000;26(3):565-81. (In eng). DOI: 10.1111/j.1728-4457.2000.00565.x.

4. Quandelacy TM, Viboud C, Charu V, Lipsitch M, Goldstein E. Age- and sex-related risk factors for influenza-associated mortality in the United States between 1997-2007. Am J Epidemiol 2014;179(2):156-67. (In eng). DOI: 10.1093/aje/kwt235.

5. CDC. Coronavirus Disease 2019 (COVID-19): Older Adults. Atlanta: Centers for Disease Control and Prevention, U.S. Department of Health & Human Services; 2020.

6. Team CC-R. Coronavirus Disease 2019 in Children - United States, February 12-April 2, 2020. MMWR Morb Mortal Wkly Rep 2020;69(14):422-426. (In eng). DOI: 10.15585/mmwr.mm6914e4.

7. Shang M, Blanton L, Brammer L, Olsen SJ, Fry AM. Influenza-Associated Pediatric Deaths in the United States, 2010-2016. Pediatrics 2018;141(4) (In eng). DOI: 10.1542/peds.2017-2918.

8. CDC. Archived Estimated Influenza Illnesses, Medical visits, Hospitalizations, and Deaths in the United States — 2017–2018 influenza season. National Center for Immunization and Respiratory Diseases (NCIRD), Centers for Disease Control and Prevention, U.S. Department of Health & Human Services; 2019.

9. Taubenberger JK, Morens DM. 1918 Influenza: the mother of all pandemics. Emerg Infect Dis 2006;12(1):15-22. (In eng). DOI: 10.3201/eid1201.050979.

10. Bautista E, Chotpitayasunondh T, Gao Z, et al. Clinical aspects of pandemic 2009 influenza A (H1N1) virus infection. N Engl J Med 2010;362(18):1708-19. (In eng). DOI: 10.1056/NEJMra1000449.

11. Guzik TJ, Mohiddin SA, Dimarco A, et al. COVID-19 and the cardiovascular system: implications for risk assessment, diagnosis, and treatment options. Cardiovasc Res 2020 (In eng). DOI: 10.1093/cvr/cvaa106.

12. Bots SH, Peters SAE, Woodward M. Sex differences in coronary heart disease and stroke mortality: a global assessment of the effect of ageing between 1980 and 2010. BMJ Glob Health 2017;2(2):e000298. (In eng). DOI: 10.1136/bmjgh-2017-000298.

13. 2016 GHE. Deaths by Cause, Age, Sex, by Country and by Region, 2000-2016. Geneva: World Health Organization; 2018.

14. Haycock PC, Burgess S, Nounu A, et al. Association Between Telomere Length and Risk of Cancer and Non-Neoplastic Diseases: A Mendelian Randomization Study. JAMA Oncol 2017;3(5):636-651. (In eng). DOI: 10.1001/jamaoncol.2016.5945.

15. Arbeev KG, Verhulst S, Steenstrup T, et al. Association of Leukocyte Telomere Length With Mortality Among Adult Participants in 3 Longitudinal Studies. JAMA Netw Open 2020;3(2):e200023. (In eng). DOI: 10.1001/jamanetworkopen.2020.0023.

16. Aviv A. Telomeres and COVID-19. FASEB J 2020 (In eng). DOI: 10.1096/fj.202001025.

17. Aviv A, Shay JW. Reflections on telomere dynamics and ageing-related diseases in humans. Philos Trans R Soc Lond B Biol Sci 2018;373(1741) (In eng). DOI: 10.1098/rstb.2016.0436.

18. Factor-Litvak P, Susser E, Kezios K, et al. Leukocyte Telomere Length in Newborns: Implications for the Role of Telomeres in Human Disease. Pediatrics 2016;137(4) (In eng). DOI: 10.1542/peds.2015-3927.

19. Morselli E, Santos RS, Criollo A, Nelson MD, Palmer BF, Clegg DJ. The effects of oestrogens and their receptors on cardiometabolic health. Nat Rev Endocrinol 2017;13(6):352-364. (In eng). DOI: 10.1038/nrendo.2017.12.

20. Brown SB, Hankinson SE. Endogenous estrogens and the risk of breast, endometrial, and ovarian cancers. Steroids 2015;99(Pt A):8-10. (In eng). DOI: 10.1016/j.steroids.2014.12.013.

21. Spagnolo PA, Manson JE, Joffe H. Sex and Gender Differences in Health: What the COVID-19 Pandemic Can Teach Us. Ann Intern Med 2020 (In eng). DOI: 10.7326/M20-1941.

22. Hall OJ, Klein SL. Progesterone-based compounds affect immune responses and susceptibility to infections at diverse mucosal sites. Mucosal Immunol 2017;10(5):1097-1107. (In eng). DOI: 10.1038/mi.2017.35.

23. Reynolds RF, Obermeyer CM. Age at natural menopause in Spain and the United States: results from the DAMES project. Am J Hum Biol 2005;17(3):331-40. (In eng). DOI: 10.1002/ajhb.20121.

24. Migeon BR. Why females are mosaics, X-chromosome inactivation, and sex differences in disease. Gend Med 2007;4(2):97-105. (In eng). DOI: 10.1016/s1550-8579(07)80024-6.

25. Libert C, Dejager L, Pinheiro I. The X chromosome in immune functions: when a chromosome makes the difference. Nat Rev Immunol 2010;10(8):594-604. (In eng). DOI: 10.1038/nri2815.

26. Devaux CA, Rolain JM, Raoult D. ACE2 receptor polymorphism: Susceptibility to SARS-CoV-2, hypertension, multi-organ failure, and COVID-19 disease outcome. J Microbiol Immunol Infect 2020;53(3):425-435. (In eng). DOI: 10.1016/j.jmii.2020.04.015.

27. Lieb W, Graf J, Götz A, et al. Association of angiotensin-converting enzyme 2 (ACE2) gene polymorphisms with parameters of left ventricular hypertrophy in men. Results of the MONICA Augsburg echocardiographic substudy. J Mol Med (Berl) 2006;84(1):88-96. (In eng). DOI: 10.1007/s00109-005-0718-5.

28. Cuspidi C, Meani S, Sala C, Valerio C, Negri F, Mancia G. Age related prevalence of severe left ventricular hypertrophy in essential hypertension: echocardiographic findings from the ETODH study. Blood Press 2012;21(3):139-45. (In eng). DOI: 10.3109/08037051.2012.668662.

29. Burrell LM, Harrap SB, Velkoska E, Patel SK. The ACE2 gene: its potential as a functional candidate for cardiovascular disease. Clin Sci (Lond) 2013;124(2):65-76. (In eng). DOI: 10.1042/CS20120269.

30. Perls TT. Male Centenarians: How and Why Are They Different from Their Female Counterparts? J Am Geriatr Soc 2017;65(9):1904-1906. (In eng). DOI: 10.1111/jgs.14978.

